# COVID-19 and Multisystem Inflammatory Syndrome in Latin American children: a multinational study

**DOI:** 10.1101/2020.08.29.20184242

**Authors:** Omar Yassef Antúnez-Montes, Maria Isabel Escamilla, Augusto Flavio Figueroa-Uribe, Erick Arteaga-Menchaca, Manuel Lavariega-Saráchaga, Perla Salcedo-Lozada, Priscilla Melchior, Rodrigo Beréa de Oliveira, Juan Carlos Tirado Caballero, Hernando Pinzon Redondo, Laura Vanessa Montes Fontalvo, Roger Hernandez, Carolina Chavez, Francisco Campos, Fadia Uribe, Olguita del Aguila, Jorge Alberto Rios Aida, Andrea Parra Buitrago, Lina Maria Betancur Londoño, León Felipe Mendoza Vega, Carolina Almeida Hernández, Michela Sali, Julian Esteban Higuita Palacio, Jessica Gomez-Vargas, Adriana Yock-Corrales, Danilo Buonsenso

**Author notes:** Contributed equally as first authors (Omar Yassef Antúnez-Montes, Maria Isabel Escamilla). Contributed equally as last authors (Adriana Yock-Corrales, Danilo Buonsenso). **Corresponding authors** Danilo Buonsenso, Largo A. Gemelli 8, 00168, Rome, Italy, For spanish-only speaking correspondences, Omar Yassef Antúnez-Montes, Ciudad de México, México, For portuguese-only speaking correspondences, Rodrigo Beréa de Oliveira, Sao Luiz Hospital, Sao Luiz, Brazil. **Funding Source** No external funding for this manuscript. **Data sharing statement** data available upon request to the corresponding author (DB). **Table of Contents Summary** This study shows a high number of severe form of COVID-19 and MIS-C in Latin American Children. **What’s Known on This Subject** Studies from China, Europe and the United States showed that COVID-19 in children is relatively milder compared with adult. However, data are lacking from Latin America despite it has been severely affected by the pandemic. **What This Study Adds** This study adds new data about pediatric COVID-19 in Latin America, describing a generally more severe form of COVID-19 and a high number of MIS-C compared with previous studies. **Contributors statement** OYAM and DB conceived of the study. OYAM, DB, MIE designed the study. OYAM, DB and MS cleaned and analyzed the data, made the figures, wrote the first draft of the manuscript. All authors actively contributed data to the study, reviewed the manuscript, and approved the final manuscript for submission.

## Abstract

**Background:** To date, there are no comprehensive data on pediatric COVID-19 from Latin America. This study aims to assess COVID-19 and Multisystem Inflammatory Syndrome (MIS-C) in Latin American children, in order to appropriately plan and allocate resources to face the pandemic on a local and International lever

**Methods:** Ambispective multicentre cohort study from five Latin American countries. Children aged 18 years or younger with microbiologically confirmed SARS-CoV-2 infection were included.

**Findings:** 409 children were included, with a median age of 53.0 years (IQR 0.6–9.0). Of these, 95 191 (23.2%) were diagnosed with MIS-C. 191 (46.7%) children were admitted to hospital and 52 (12.7%) required admission to a Pediatric Intensive Care Unite (PICU). 92 (22.5%) patients required oxygen support: 8 (2%) were started on continuous positive airway pressure (CPAP) and 29 (7%) on mechanical ventilation. 35 (8.5%) patients required inotropic support. The following factors were associated with PICU admission: pre-existing medical condition (P < 0.0001), immunodeficiency (P = 0.01), lower respiratory tract infection (P< 0.0001), gastrointestinal symptoms (P = 0.006), radiological changes suggestive of pneumonia and acute respiratory distress syndrome (P< 0.0001), low socioeconomic conditions (P 0.009).

**Conclusions:** This study shows a generally more severe form of COVID-19 and a high number of MIS-C in Latin American children, compared with studies from China, Europe and North America, and support current evidence of a more severe disease in Latin/Hyspanic children or in people of lower socioeconomic level. The findings highlight an urgent need of more data of COVID-19 in South America.

## Introduction

Despite several months having passed since the first description of the SARS-CoV-2 outbreak in China, twenty millions of cases reported worldwide and thousands of deaths, many questions about the COVID-19 pandemic have no answers yet. In particular, the puzzle of the impact of COVID-19 in children still has a number of missing pieces. Initial data from China, Italy and the United States (US) gave optimistic data with a limited number of children involved by the pandemic and rare complications (1). The first multinational study from Europe including more than 500 children from a network of pediatric infectious diseases and pulmunology specialists from major centers confirmed a relatively milder disease compared with adults (2). However, later during the pandemic, several authors from Europe and the US reported an unusual rate of multisystem inflammatory syndromes without a known etiology and temporally related to SARS-CoV-2 (3-5). This syndrome is now described as MIS-C (or Pediatric Multisystem Inflammatory Syndrome temporally related to SARS-CoV-2 – PIMS-TS) and, although rare, is associated with a non negligible number of Pediatric Intensive Care Unit (PICU) admissions and death (3-5). This more severe form of COVID-19 has been described more frequently in specific ethinic categories of children, in particular black, afro-caribbean and latino/hispanic children 6). However, although South America is currently severely involved by the COVID-19 pandemic, multinational studies from this area, similarly to those described in Europe and US, are missing, with the exception of two PICU studies from Chile (7) and Brasil (8), traditionally the two Latin American countries more involved in research projects. In order to provide fast and up-to-date key data on COVID-19 in children in Latin America, one of the largest and more populated areas in the world, inspired by a previous European study, we used a newly established research network (9), predominantly comprising paediatric infectious diseases specialists. Moreover, Latin America is characterized by a specific geo-political and ethnic situation and, considering the potential impact of political approaches on the pandemic, data from this area are necessary to appropriately plan and allocate resources to face the local (and indirectly the international) situation of COVID-19 pandemic, and to better understand the real impact of COVID-19 on children.

## Methods

### Study design and participants

Independent Pediatricians, Pediatric Infectious Diseases specialists and Emergency Physicians from Mexico, Colombia, Peru, Costa Rica and Brazil developed a “CoviD in sOuth aMerIcaN children – study GrOup (CoviDOMINGO)” (9) and collected cases of confirmed pediatric SARS-CoV-2 infections evaluated before or during the study period. All patients ≤ 18 years old with positive RT-PCR on at least one clinical sample (nasopharyngeal swab, bronchoalveolar lavage, blood, stool, or cerebrospinal fluid), or fulfilling the criteria for MIS-C (PIMS-TS) with microbiological documentation of SARS-CoV-2 exposure (PCR or IgG), according to the Centers for Disease and Control (CDC) were included in the study.

Data were collected on Excel spreadsheets completed by each collaborator and sent to two study core group members via email (DB and OYAM), without including personal or identifiable data. Data collection began on July 1st and was concluded on August 11th, 2020. The study was reviewed and approved by the CoviDOMINGO core group and approved by the Ethic Committee of the coordinating center and by each participating center (Mexico: COMINVETICA-30072020-CEI0100120160207; Colombia: PE-CEI-FT-06; Perù: N° 42-IETSI-ESSALUD-2020; Costa Rica: CEC-HNN-243-2020). The study was conducted in accordance with the Declaration of Helsinki and its amendments. No personal nor identifiable data were collected during the conduct of this study.

Variables collected include age, gender, symptoms, socioeconomic status, need for hospital and NICU/PICU admission, respiratory and cardiovascular support, other viral co-infections, drugs used to treat COVID-19, outcome. Respiratory symptoms were classified as upper respiratory tract infection (e.g. rhinitis, pharyngitis, tonsillitis, otitis) and lower respiratory tract infection (e.g. pneumonia, bronchitis) with or without using radiological imaging according to local guidelines and evaluating clinician’s decisions. Fever was defined as body temperature ≥ 38.0°C. The socioeconomic status was classified according to the Colombian definition of “current legal minimum wage” that is of 980.657 pesos colombianos (258.664 US dollars) and adapted to each of the other participating countries. Starting from the legal minimum wage, we classified “very low status” is the income was less than one minimum wage, “low” if equal to one wege, “medium-low” if between two and five current legal mínimum wage, “medium” if between five and eight minimum wage, “medium-high” between eight and sixteen minimum wage, “high” if more than sixteen minimum wage.

Moreover, a further section regarding MIS-C (or PIMS-Ts) has been developed. MIS-C criteria were those highlighted by the Centers for Disease Control and Prevention (https://www.cdc.gov/mis-c/hcp/). In case of MIS-C diagnoses, data regarding the organs involved, therapeutic strategies and outcome were analyzed.

Collected data have been made similar to previous multinational studies from other countries (2) to allow to establish if different epidemiological/ethinic settings were associated with different characteristics of pediatric COVID-19, since recent evidences are suggesting that genetic factors may predispose to more severe forms of COVID-19 or to the development of MIS-C (PIMS-TS).

### Statistical analyses

Data were analyzed using SPSS (SPSS, Chicago, IL). Differences in frequencies were evaluated by the Fisher exact test. The nonparametric Mann-Whitney U test was used to compare medians for unpaired comparisons and the Wilcoxon test for paired comparisons, and the Kruskal-Wallis test was used to compare medians among the different groups. Differences were considered significant at P values of 0.05.

### Translations

A supplementary material file is uploaded with translation of the manuscript in Spanish and Portugues, to allow non English speakers/readers to be able to access the information of this study. The translation have been performed by native speakers authors of this study: OYAM and MIE (Spanish), PM and RBdO (Portugues).

## Results

SARS-CoV-2 infection were reported from 14 health-care institutions in 5 Latin America countries: Mexico, Colombia, Perù, Costa Rica, Brasil (figure 1a). Colleagues from Venezuela agreed to participate but did not receive authorization to share COVID-19 cases by their Institution, while colleagues from Ecuador declared to be overwhelmed by workload and have not been able to upload their cases for this first study. Two colleagues from Brazil experienced organizational delays in obtaining Ethic Committee approval and therefore did not included their cases and were not even allowed to submit gross numbers. 409 children with PCR-confirmed SARS-CoV-2 infection were included in the final analyses. Of these, 95 (23.2%) fulfilled CDC criteria for MIS-C (figure 1b).

**Figure 1.**
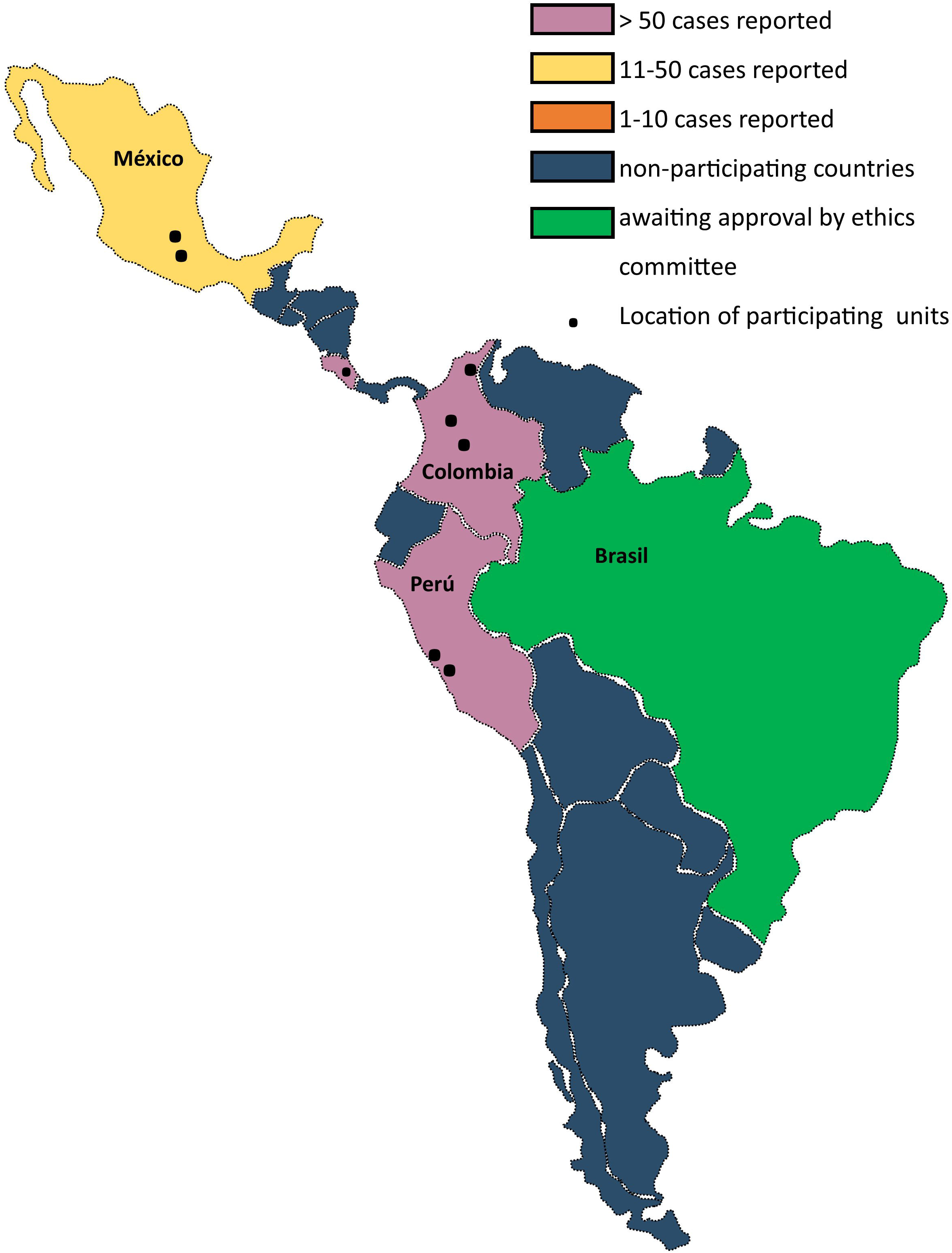
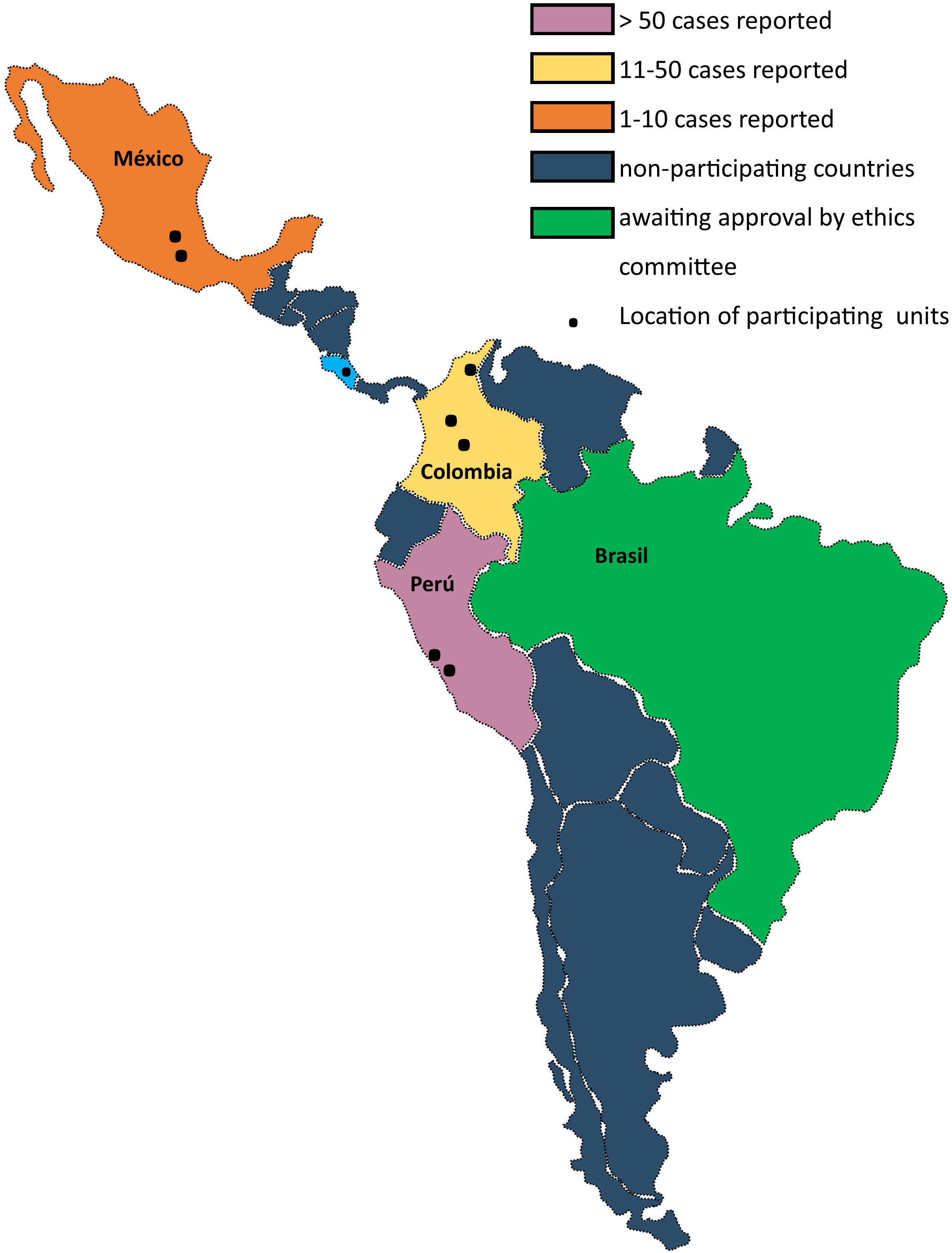
Location of participating units and number of paediatric COVID-19 (A) and Multisystem Inflammatory Syndrome (B) cases reported by country

The median age of the study population was 3.0 years (IQR 0.6–9.0), ranging from 2 days to 18 years (table 1). 117 (28.6%) participants younger than 12 months. 222 children were male (54.3%). The most common source of infection was a parent, considered the index case in 165 (40.3%) cases; for 5 (1.2%) individuals, the most probable index case was a sibling. In 62 (15.2%) individuals, the index case was a person outside of the immediate family, while it was unknown in 177 cases (43.3%). 191 (46.7%) children were admitted to hospital and 52 (12.7%) required admission to a Pediatric Intensive Care Unite (PICU), corresponding to 27% of those admitted to hospital.

**Table 1.** Characteristics of the study population and by requirement of PICU admission. Pediatric Intensive Care Unit (PICU); Acute Respiratory Distress Syndrome (ARDS); Multisystem Inflammatory Syndrome (MIS-C);

|  | Entire cohort<br>(n= 409) | Not admitted to<br>PICU(n=357) | Admitted to<br>PICU (n= 52) | p value |
| --- | --- | --- | --- | --- |
| **Age, years** |  |  |  | > 0.05 |
| <1 month | 36 (8.8) | 29 (8.1) | 7 (13.5) |  |
| 1-23 months | 163 (40) | 145 (40.6) | 18 (34.6) |  |
| 2–5 | 54 (13.3) | 48 (13.4) | 6 (11.5) |  |
| 5–10 | 63 (15.5) | 51 (14.3) | 12 (23.1) |  |
| >10 | 91 (22.4) | 83 (23.3) | 8 (15.4) |  |
| missing | 2 (0.5) | 1 (0.3) | 1 (1.9) |  |
| **Sex** |  |  |  | > 0.05 |
| Female | 187 (45.7) | 165 (46.2) | 22 (42.3) |  |
| Male | 222 (54.3) | 192 (53.8) | 30 (57.7) |  |
| **Index Case** |  |  |  | 0.003 |
| Parent | 165 (40.3) | 152 (42.6) | 13 (25) |  |
| Sibling | 5 (1.2) | 4 (1.1) | 1 (1.9) |  |
| Other | 62 (15.2) | 58 (16.2) | 4 (7.7) |  |
| Unknown | 177 (43.3) | 143 (40.1) | 34 (65.4) |  |
| **Pre-existing medical conditions** | 83 (61.2) | 60 (16.8) | 23 (44.2) | < 0.0001 |
| Congenital Heart Disease | 2 (0.5) | 2 (0.5) | - |  |
| Congenital Syndromes | 6 (1.5) | 1 (0.3) | 5 (9.6) |  |
| Immunological Diseases | 7 (1.7) | 3 (0.8) | 4 (7.7) |  |
| Neurological Disorders | 10 (2.5) | 9 (2.5) | 1 (1.9) |  |
| Others | 58 (14.3) | 45 (12.6) | 13 (25) |  |
| Immunosuppressive therapy | 12 (2.9) | 7 (19.6) | 5 (9.6) | 0.002 |
| Known immunodeficiency | 18 (4.4) | 12 (3.3) | 6 (11.5) | 0.008 |
| Chemotherapy in past | 14 (3.4) | 10 (2.8) | 4 (7.7) | > 0.05 |
| **Signs and symptoms** |  |  |  |  |
| Asymptomatic | 49 (12) | 49 (13.7) | - |  |
| Pyrexia | 238 (58) | 201 (53.6) | 34 (65.4) | 0.052 |
| Upper respiratory tract infection | 244 (60) | 222 (62.2) | 22 (42.3) | 0.005 |
| Lower respiratory tract infection | 102 (25) | 64 (17.9) | 38 (73.1) | < 0.0001 |
| Gastrointestinal | 101 (24.7) | 80 (22.4) | 21 (40.4) | 0.006 |
| Headache | 48 (11.7) | 40 (11.2) | 8 (15.4) | > 0.05 |
| **Radiological findings** |  |  |  |  |
| Suggestive of pneumonia | 170 (41.5) | 127 (35.6) | 43 (82.7) | < 0.0001 |
| Suggestive of ARDS | 17 (4) | 5 (1.4) | 12 (23.1) | < 0.0001 |
| Viral co-infection | 14 (3.4) | 10 (2.8) | 4 (7.7) | > 0.05 |
| **MIS-C** | 95 (23) | 75 (21) | 20 (38.5) | 0.004 |
| **Socioeconomic status** |  |  |  | 0.009 |
| Very low | 67 (16.4) | 51 (14.3) | 16 (30.7) |  |
| Low | 82 (20) | 76 (21.3) | 6 (11.5) |  |
| Low-medium | 144 (35.2) | 123 (34.4) | 21 (40.4) |  |
| Medium | 40 (9.8) | 38 (10.6) | 2 (3.8) |  |
| Medium-high | - | - | - |  |
| High | - | - | - |  |
| Unknown | 76 (18.6) | 66 | 10 |  |
| **Deaths** | 17 (4.2) | - | 17 |  |

Fever was reported in 238 cases (58%) children. 244 (60%) children had symptoms of upper respiratory tract infection and 102 (25%) children had lower respiratory tract symptoms. 101 (24.7%) had diarrhea. 53 (13%) individuals were asymptomatic. 77 (18.8) children had pre-existing comorbidities, 18 (4.4%) had pre-existing immunological disorders. 49 (12%) were asymptomatic.

A chest x-ray was done in 157 (38.4%) patients. Of those, 84 had signs of COVID-19 pneumonia, with 33 (21%) having consolidations and 51 (33%) interstitial disease.

In 13 (3.2%) children, a viral co-infection was detected (1 rhinovirus, 1 Epstein-Barr Virus, 1 respiratory syncytial virus, 1 Influenza A, the others not reported), in one more case a co-infection with *M. pneumoniae* was detected. Co-infections were not associated with a higher risk of PICU admission nor invasive respiratory support.

317 (77%) individuals did not require respiratory support. 92 (22.5%) patients required oxygen support: 8 (2%) were started on continuous positive airway pressure (CPAP) and 29 (7%) on mechanical ventilation. One child received extracorporeal membrane oxygenation. 35 (8.5%) patients required inotropic support. The following factors were significantly associated with PICU admission and need of invasive ventilation: pre-existing medical condition (P < 0.0001), known immunodeficiency (P = 0.01 and P = 0.006, respectively), lower respiratory tract infection (P< 0.0001), gastrointestinal symptoms (only associated with PICU admission, P = 0.006), radiological changes suggestive of pneumonia and acute respiratory distress syndrome (ARDS) (P< 0.0001). Also, low socioeconomic conditions were associated with PICU admission (P 0.009) and mechanical ventilation (P< 0.04). An unknown exposure to index-case was associated with PICU admission (P 0.003). In multivariable analysis, the factors, with the exception of radiological changes suggestive of ARDS, remained associated with PICU admission.

Drugs with possible or known antiviral activities were rarely used: hydroxychloroquine (10 cases, 2.4%), oseltamivir (9 cases, 2.2%), lopinavir–ritonavir (5 cases, 1.2%), chloroquine (one case, < 1%), while remdesivir, favipiravir, zanamivir and ribavirin were never used. Regarding immunomodulatory medication, 63 (15%) patients received systemic corticosteroids, 40 (10%) intravenous immunoglobulin, two (<1%) received tocilizumab.

Seventeen patients (4.2%) died (median age 1 year, min 5 days max 16 years) died. Of these, two were newborns and five were infants younger than 12 months of age. Fatal cases were described in Mexico (6), Colombia (5), Perù (6).

### Multisystem inflammatory syndrome (MIS-C)

As of August 11, 2020, a total of 95 MIS-C patients (23%) had been reported in our cohort (Figure 2). The median patient age was 7 years (range = 1 month–17 years); 25 (54.7%) were male, all being mestizos as ethnic group (Latin living in South America). Seven were defined as extremely poor, five as class 2, sixty-four as class 3, eighteen as class 4 of socioeconomic background according to the local classifications (Table 2).

**Figure 2.**
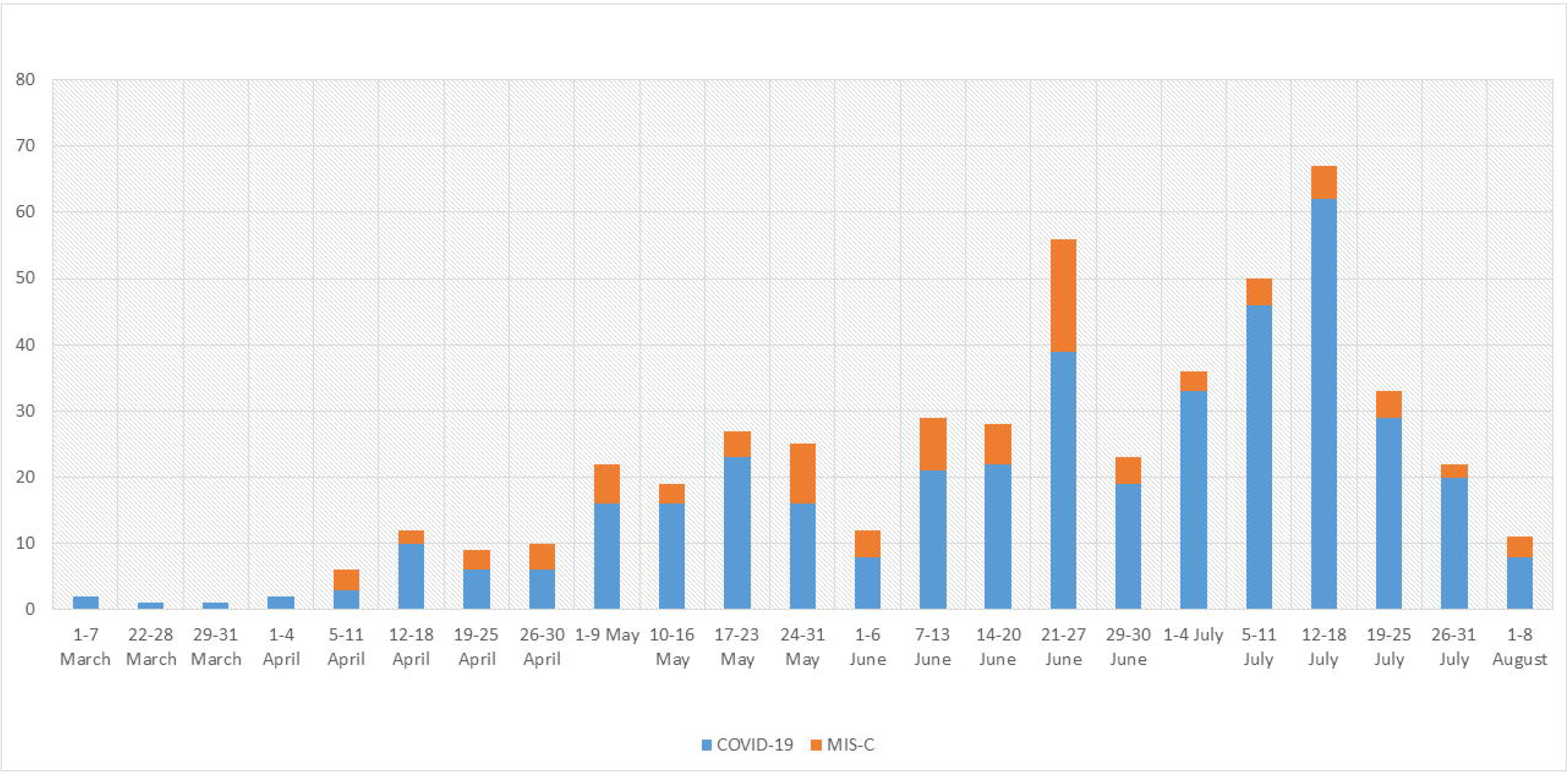
Weekly cumulative cases of Multisystem Inflammatory Syndrome compared with COVID-19

**Table 2.**
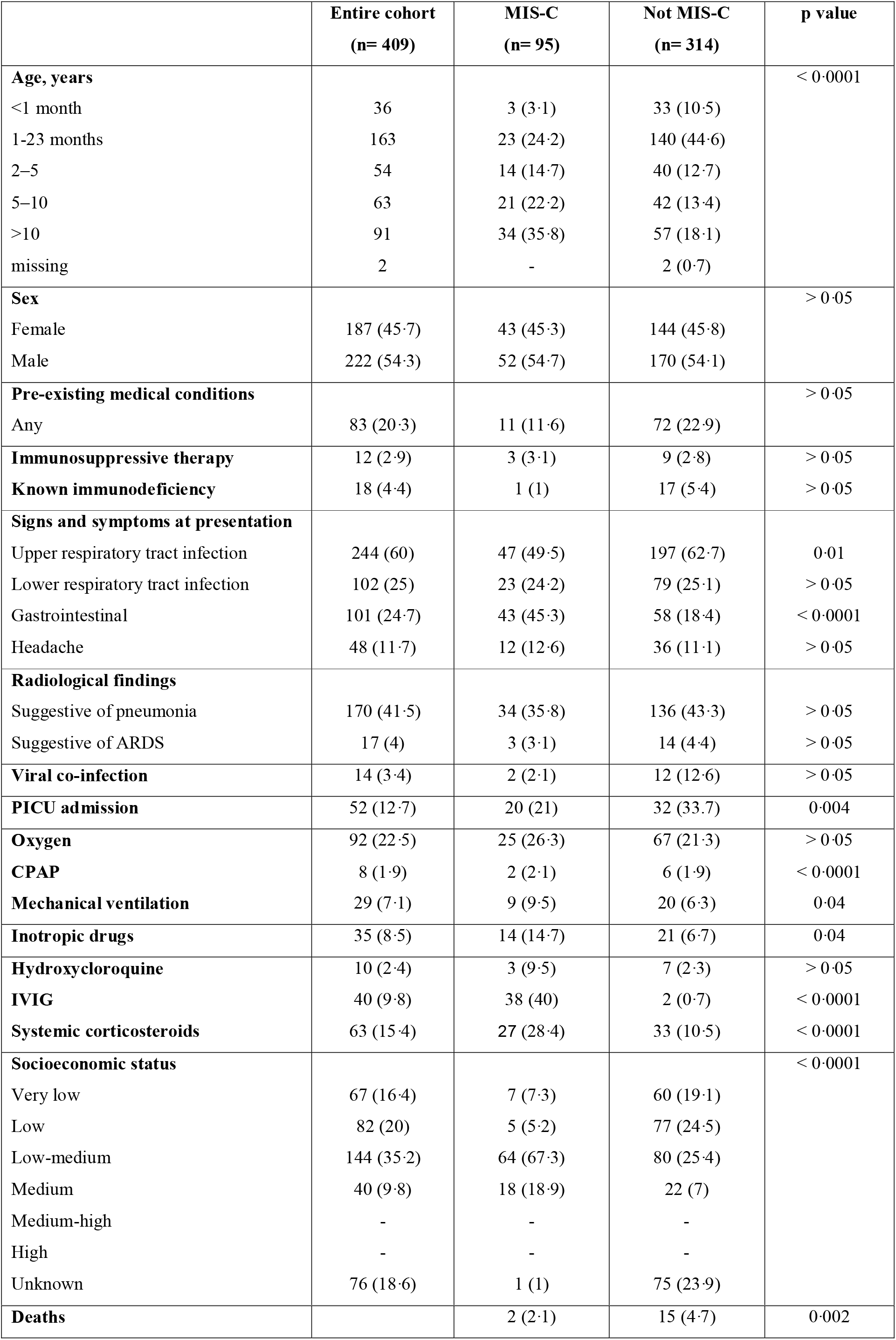
Characteristics of the study population according to Multisystem Inflammatory Syndrome (MIS-C) diagnosis. Pediatric Intensive Care Unit (PICU); Acute Respiratory Distress Syndrome (ARDS); Continuous Positive Airway Preassure (CPAP)

Eleven children had a preexisting underlying medical conditions before MIS-C onset (11.6%). 43 (45.3%) patients had gastrointestinal symptoms, eleven (11.5%) cardiovascular involvement (of which five developed coronary dilatation, four pericardial effusion, two myocarditis) and 14 children (14.7%) required inotropic support for shock, seven (7.3%) had joint symptoms. Twenty (21%) were admitted to PICU. Two children (2.1%) died. Mortality was higher in the non MIS-C group.

Of the 95 MIS-C patients, all had evidence of SARS-CoV-2 infection. Serology was performed in 88 cases and resulted positive in 72 (81.8%), those with negative or not performed serology had a positive nasopharyngeal swab.

The following factors were significantly associated with a diagnosis of MIS-C: older age (P < 0.0001), gastrointestinal symptoms (P < 0.0001), a lower socioeconomic status (P < 0.0001), higher use of inotropic agents (P 0.04), intravenous immunoglobulin (IVIG) and steroids (P < 0.0001).

38 (40%) received (IVIG), 27 (28.4%) steroids, 14(14.7%) were treated with inotropic agents and 2 (2.1%) received tolicizumab. Moreover, 3 children (3.1) received hydroxycloroquine.

## Discussion

In this study, we report data on the first multinational, multicentre study of pediatric COVID-19 in Latin America, specifically from countries that did not reported yet detailed data about COVID-19 in children. Although several studies about pediatric COVID-19 have been published from China, Europe and US, a comprehensive picture from South America was still missing and in this report we highlight some important differences from other centers with potential public health implications. In fact, although the most common symptoms (fever, respiratory symptoms and diarrhea) were similarly described compared to other studies, and a lower proportion of children required hospital admission compared to a European study (46.7% vs 62%, respectively), a larger number of Latin American children required PICU admission (12.7% vs 8%) and, importantly, seventeen children (4.2%) died (compared with 4 (0.68%) of the European cohort) (6). Also, the median age of the dead children in Latin America was much lower than every described study so far, with seven children younger than one year of age. Although the participating centers were mainly referral national Hospitals and these data represent, therefore, the more severe spectrum of COVID-19 in Latin America, the same type of centers were enrolled by Goezinger et al (6), suggesting that the differences are real and that the SARS-CoV-2 is having a stronger impact in South America. Of note, all the authors involved in this study are directly involved on the front-line and report a growing number of cases while the described data have been analysed, since most South American countries are in the middle of the COVID-19 peak. The higher number of severe disease and deaths reported in our series, however, is not completely unexpected, since acute COVID-19 has been reported to disproportionately affect Hispanics and blacks (10). Long-standing inequities in the social determinants of health, such as housing, economic instability, insurance status, and work circumstances of patients and their family members have systematically placed social, racial, and ethnic minority populations at higher risk for COVID-19 and more severe illness (6), and our data further support this view. In fact, the epidemiological context of our cohort is different from previous large studies (1,2, 11-14): a household transmission of the infection was documented only in 170 (41.5%) cases and the index-case remained unknown in 177 cases (43.3%). This may reflect different political decisions since in these areas a strict lockdown was not established, allowing a wider community spread of SARS-CoV-2 and higher risk for children to be infected during common daily activities. In adjunction, the family socioeconomic impact have a clear impact in our study: 149 families (36.4%) did not earn more than the current legal minimum wage (258 US dollars). Lower socioeconomic conditions were significantly associated with need of PICU admission or mechanical ventilation (P< 0.0001) and most of those who died were classified as low-very low socioeconomic conditions. Notably, none of the children classified in our study were of high socioeconomic level, although this might be due to preference of those families to be evaluated in private centers.

In our cohort, the risk factors associated with need of PICU admission were similar to those described in the multinational European study (presence of respiratory symptoms or radiologic evidence of COVID-19 pneumonia/ARDS, pre-existing medical conditions, immunological conditions of immune suppressive therapies) (2). Conversely, viral co-infections were rarely detected in our cohort and not associated with a more severe disease, possibly due to the seasonal period of our study.

Drugs with possible antiviral activity (including hydroxychloroquine) were rarely used, compared with other studies (2, 11-13), while intravenous steroids were commonly administered. This is possibly related to the later arrival of the COVID-19 peak in South America, when data about the low benefit of hydroxychloroquine were reported (14, 15) and dexamethasone showed greater benefits in the RECOVERY trial (16)

In our cohort, a high number of children were diagnosed with MIS-C (PIMS-TS). Since the case definition is nonspecific and confirmatory laboratory testing does not exist, it can be difficult to distinguish MIS-C from other systemic inflammatory conditions such as severe acute COVID-19 and Kawasaki disease (17). For this reason, the number of MIS-C can be overestimated. For example, as the COVID-19 pandemic spreads, and more children are exposed to SARS-CoV-2 with subsequent seroconversion, patients with systemic inflammatory diseases (not only Kawasaki Diseases) might be erroneously as MIS-C because of an incidental finding of antibodies to SARS-CoV-2. However, since previous reports described a higher incidence of this condition in latin/hyspanic children (6), this number can also represent a real higher incidence of MIS-C in Latin America. Currently, there are no other multinational studies from this area to confirm our data, however an ongoing national collection from Chile reported, as of August 18th, a total 149 MIS-C (https://mobile.twitter.com/jptorrest/status/1295136584199737346) and a national study from Brasil reported 79 children requiring PICU admission with 10 of them classified as MIS-C (8).

Overall, the age distribution of the patients in this analysis is similar to previously published studies (median age 7), confirming the older age of this group of children compared with Kawasaki Disease. Differently from a recent US report (6), however, the age range of our cohort was much higher, including very young infants aw well. 2.1% died, a slightly higher proportion compared with the US cohort (1.8%), highlighting again that both genetic and socioeconomic factors may contribute to a higher proportion of COVID-19 severity in Latin America (10).

Our study has some limitations to address. The main limitation of this study relates to the variables collected. As happened during a multinational European study, this one was performed during the Latin American peak with clinicians struggling in the front-line, usually with limited human resources to dedicate extra time for clinical research. For example, detailed blood tests were not collected. However, at this time of the pandemic enough laboratory data on pediatric COVID-19 have been published and we think that a first, large, multinational picture of SARS-CoV-2 infection in Latin American children was more important than smaller, more detailed studies. Also, the different centers may have used different decision rules to perform SARS-CoV-2 test in children. Another limitation concerns MIS-C cases. Since MIS-C is a clinical diagnosis with no confirmatory test, and that the CDC case definition is broad, some cases may have been misdiagnosed and, therefore, the real MIS-C cases being lower or higher. For example, some severe cases of acute COVID-19 may overlap with MIS-C. Also, some details about MIS-C were not included in our data collection, including the possible skin, renal and neurological involvement during MIS-C. Despite these limitations, this study provides the most comprehensive overview on COVID-19 in South American children to date.

In conclusion, our study adds new data about the Latin American face of the pediatric SARS-CoV-2 pandemic, describing a generally more severe form of COVID-19 and a high number of MIS-C compared with studies from China, Europe and North America. Unfortunately, a significant number of children in our cohort required PICU admission and a non-negligible number of deaths have been reported. These data support current evidence of a more severe disease in Latin/Hyspanic children. Importantly, a clear association between more severe disease and socioeconomic status have been found, supporting worldwide evidence of the unequal impact of COVID-19 on fragile people (18) and highlighting its impact on children as well.

Our study further supports the importance of continuous monitoring of the impact of COVID-19 and MIS-C in children, and the need of actively including Low-Middle Income Countries or areas with political instability in collaborative studies, in order to have a better knowledge of the real burden of COVID-19 in children. Hopefully, this study will open the road to wider collaborations and the beginning of a comprehensive multinational study including all countries from Latin America.

## Data Availability

available upon request

## Acknowledgements

We are grateful to all collaborators that help the development of the DOMINGO study group that, for different reasons, were not able to contribute with data: Rosa Arana Sunohara (Perù), Jorge Omar Flores del Razo (Mexico), Jaime Amadeo Tasayco Muñoz (Perù), Teresa ochoa (Perù), Lara Limansky (argentina), Grimaldo de los Angeles Ramírez Cortez (Perù)

## Abbreviations

MIS-C: Multisystem Inflammatory Syndrome
PIMS-TS: Pediatric Multisystem Inflammatory Syndrome temporally related to SARS-CoV-2
PICU: Pediatric Intensive Care Unit
CDC: Centers for Disease and Control

